# Availability and Quality of Antenatal and Pediatric Clinical Services at Primary Healthcare Facilities in Western Kenya: An Application of the Primary Care Assessment Tool

**DOI:** 10.64898/2025.12.10.25342014

**Authors:** Sarah Hawi Ngere, Kitiezo Aggrey Igunza, Edwin Kiplelgo, Meghna Ray, Namrata Ramakrishna, Maria Velez, Kennedy Ochola, Eucabeth Awuonda, Anne Aluoch, Alex Chweya, Abraham Otieno, Harun Owuor, Christopher Mugah, Peter Otieno, Richard Omore, Dickens Onyango, Cynthia G. Whitney, Portia C. Mutevedzi, Hellen Muttai, Victor Akelo, Chris A Rees

## Abstract

Child mortality remains unacceptably high in Western Kenya and >60% of children who die access primary healthcare facilities in the immediate period prior to their death. This study aimed to assess the availability and quality of antenatal and pediatric clinical services at primary healthcare facilities within the catchment area of an ongoing childhood mortality surveillance program. This study employed a cross-sectional design using the standardized Primary Care Assessment Tool (PCAT) in Siaya and Kisumu counties in Western Kenya. Ten primary healthcare facilities were randomly selected in each county. The PCAT was administered to clinicians and caregivers of pediatric patients via REDCap. Qualitative responses were reviewed, coded, and analyzed thematically to complement quantitative findings. A total of 57 healthcare providers and 99 caregivers participated in the survey. Most healthcare providers were female (77.2%), with a median age of 36 years; one-third were bedside nurses (33.3%) and nearly one-third were clinical officers (31.6%). Most caregivers were mothers (90.9%), female (93.9%), with a median age of 27 years. Nearly half of caregivers (47.4%) reported waiting less than 30 minutes for services, but most perceived limited availability of after-hours care. Delivery services (64.6%), breastfeeding support (63.6%), and postpartum depression support (61.6%) were less available compared to other services. Continuity of care was limited, with only 31.6% of providers stating they “definitely” knew their patients’ lives. While referral letters were commonly issued (91.2%), less than half of providers (45.6%) reported that transportation support was available for referrals. Primary healthcare services in Western Kenya were generally accessible for pediatric and antenatal care but limited for postnatal care. Referral systems suffered from suboptimal coordination and transportation. Strengthening postnatal service delivery, improving availability of ambulances, and enhancing knowledge about patients are urgent priorities for improving care quality and availability.

## Introduction

From the 1990s to the 2020s, there was a dramatic reduction in childhood mortality rates globally.(1) These gains resulted partly from expanded treatment availability, improvements in country-wide and individual socioeconomic status, and widespread vaccination programs.(2) However, progress in the global reduction in childhood mortality has not been equitably distributed with nearly 60% of deaths among children aged <5 years observed to occur in sub-Saharan Africa alone. In Western Kenya, it is estimated that mortality rates among children aged <5 years are as high as 72 per 1,000 live births.(3)

Results from previous studies suggest that most childhood deaths are preventable through access to, and the provision of, timely and high-quality clinical care.(4,5) Some of these studies have shown patient-level factors to be associated with childhood mortality suggesting the need for increased access to primary healthcare facilities as an important effort to potentially reduce childhood mortality further.(6–8) While these factors are crucial to childhood mortality reduction, the availability and quality of clinical care children receive when they reach healthcare facilities in sub-Saharan Africa has less frequently been studied, and where data exist, substantial gaps in referrals and timely access to clinical care in primary healthcare facilities have been reported.(9) Understanding possible gaps in primary healthcare access is important to inform evidence-based and feasible interventions needed to strengthen health systems to avert further childhood mortality.

Previous research on health facility capacity has identified the need for a systems-level perspective on quality of care.(11–13) The Primary Care Assessment Tool (PCAT) is a validated series of surveys designed to identify health system-level facilitators and barriers to delivering high quality clinical care in primary healthcare facilities.(14,15) The PCAT has been used to evaluate primary healthcare services in Brazil, Thailand, South Korea, Vietnam, Malawi, and South Africa.(14–18) Such an assessment has not been conducted in Kenya for pediatric and maternal health, which, as a country, has prioritized primary healthcare as part of its goal to achieve universal healthcare coverage.(19) The PCAT could provide valuable insights into system-level barriers and facilitators to high-quality care, which is critical for strengthening primary healthcare and achieving Sustainable Development Goals (SDG) aimed at ending preventable child deaths. Furthermore, PCAT has not been conducted in a targeted fashion to child health in regions with high rates of childhood mortality. Understanding the availability and quality of child and maternal health services available in primary healthcare facilities is essential, because a prospective childhood mortality surveillance program in Western Kenya determined that >60% of deceased young children accessed primary healthcare facilities in the immediate period prior to their death,(10) which suggests this may represent a crucial point for intervention to prevent childhood mortality.

The objective of this study was to assess the availability and quality of antenatal and pediatric clinical services at primary healthcare facilities within the catchment area of an ongoing childhood mortality surveillance program in Western Kenya.

## Materials and Methods

### Study Design and Setting

This study employed a cross-sectional descriptive design using the PCAT to assess the quality and availability of antenatal and pediatric healthcare services at outpatient primary healthcare facilities. Surveys were conducted between 24 October 2024 and 24 January 2025.

This study was conducted in Siaya and Kisumu counties in Western Kenya as part of Child Health and Mortality Prevention Surveillance (CHAMPS). The full procedures of CHAMPS have been described previously.(22,23) Briefly, CHAMPS aims to determine and understand causes of death to inform public health interventions and reduce future deaths among young children. To do so, CHAMPS conducts active surveillance in healthcare facilities and communities to identify stillbirths and children aged <5 years who die. For those deaths with consenting caregivers, postmortem testing is conducted using minimally invasive tissue sampling, examination of tissues, and microbiologic assays. Then, expert panels review all available data and test results to determine causes of death and make recommendations on preventability of childhood deaths. The motivation for the current study stemmed from both the frequency of primary healthcare encounters observed just before death occurred as well as the recommendations from expert panels on preventability through timely access to high quality clinical care.(4,10)

In both Siaya and Kisumu counties, level 2 and 3 primary healthcare facilities, dispensaries and health centers, respectively were selected to participate in the survey. In Kenya, health centers are those in which promotive, preventive, and curative services are provided to pregnant mothers, children, and to adult patients. Dispensaries are basic healthcare facilities that provide primary care for simple ailments and are run by nurses with supervision from nearby primary healthcare facilities.

Surveys were conducted after randomly selecting a sample of 20 of all 62 primary healthcare facilities in the CHAMPS-Kenya catchment area 10 in Kisumu County and 10 in Siaya county. Primary healthcare facilities were selected using a random number generator regardless of whether they were private or public health facilities. The selected health facilities provided a relative representation of the availability of primary healthcare services, including both public (n=43) and private (n=19) healthcare facilities in these counties.

### Survey Tool

The PCAT, a standardized validated survey tool with both quantitative and qualitative questions, was adapted to reflect the context of Western Kenya, with a specific focus on antenatal and pediatric outpatient clinical care within the CHAMPS surveillance regions. The PCAT assesses quality across several core domains that represents essential attributes of effective primary care. These domains include access to care, first contact for clinical care, ongoing clinical care, comprehensiveness of clinical care, coordination of clinical care, the extent to which clinical care is family-focused, community orientation of the clinical care provided within a facility, and cultural competency of healthcare providers.(15,18,23) The tool consists of multiple items within each domain, rated on a four-point Likert scale ranging from “definitely not” to “definitely.” Domain scores were generated by averaging item responses, and an overall primary care score was computed by aggregating the core domain scores. Higher scores reflect stronger primary care performance and better perceived quality of services. The PCAT’s structured approach provides a systematic evaluation of how well primary care facilities deliver accessible, continuous, coordinated, comprehensive, and culturally competent care

In this study, we used the 1) consumer/patient and 2) provider PCAT to evaluate primary healthcare delivery in CHAMPS catchment areas in Kenya related to antenatal and pediatric clinical care in primary healthcare facilities.

### Participants

Caregivers were eligible for inclusion if they met the following inclusion criteria: willingness to participate and provide informed consent; being a parent of a child aged <5 years or an expecting mother; considering the health facility as their primary source of care and child; and having visited the health facility at least three times. The study team used the PCAT to interview ≥3 clinicians and ≥5 caregivers of children aged <5 years at each selected healthcare facility. Since many health facilities had around three healthcare providers, all willing to participate were included in the study. A consecutive sampling technique was utilized for caregivers, whereby the first five eligible caregivers on the day the study staff visited the facility were approached for enrollment. Healthcare providers were selected based on their willingness to participate and provide informed consent, and their role in delivering clinical care to pregnant women and children at the included primary healthcare facilities.

### Study Procedures

Study staff were trained on the PCAT and were fluent in English, Dholuo, and Kiswahili.

The study employed consecutive random sampling to select participants. At the primary healthcare facilities, caregivers and expectant mothers who visited facilities for clinical services were approached to assess eligibility. Following informed consent, participants were interviewed in a quiet private room, using the appropriate PCAT form. The interviews were conducted in Dholuo, Swahili, and English depending on the comfort of the participant. The PCAT was administered via REDCap by trained CHAMPS staff to both clinicians and caregivers of pediatric patients at each participating healthcare facility. Data quality was ensured through frequent checks during the period of data collection by a data quality manager who was not involved in the data collection.

### Data analysis

We conducted descriptive statistics for frequencies and proportions using R software, version 4.5.0. For questions in the PCAT that used Likert scales, we categorized responses and calculated proportions of each response. Qualitative data were extracted from texts entries in RedCap and entered into an Excel sheet. Data was thereafter coded and subsequently reviewed and thematically analyzed. Common themes were identified and categorized through an iterative process that involved regular consultation between the first and second authors during the analysis period. The qualitative data was used to provide additional insights into data collected through quantitative methods. Qualitative themes were reported in line with quantitative responses where appropriate.

### Ethical consideration

The study was approved by the Kenya Medical Research Institute Scientific Ethics Review Unit (approval number 4876), National Commission for Science, Technology, and Innovation in Kenya (approval number 24/34659), and the Jaramogi Oginga Odinga Teaching and Referral Hospital Ethics Research Committee. A written informed consent was provided to all respondents, all of whom were literate. All pertinent information about the study to guide them in making informed decisions on whether to participate or not.

## Results

### Demographic Characteristics

The team approached 67 healthcare providers and 174 caregivers for participation. Of these, 57 (85.0%) healthcare providers and 99 (56.9%) caregivers responded to the survey at 20 primary healthcare facilities.

Among the 57 healthcare providers who completed the survey, the majority were female (77.2%), median age was 36 years (interquartile range [IQR] 31, 43 years), 33.3% were bedside nurses, and 31.6% were clinical officers (Table 1). Despite reporting a median of 12 (IQR 6, 20) consultations with children per day, only 22.8% of healthcare providers had received prior specialty training in pediatric care.

**Table 1.** Demographics of surveyed healthcare providers at 20 healthcare facilities in Western Kenya (N=57)

|  | n (%) |
| --- | --- |
| **Sex** |  |
| Male | 13 (22.8) |
| Female | 44 (77.2) |
| **Age, median (interquartile range [IQR])** | 36 (31, 43) |
| **Role** |  |
| Specialist/Consultant Doctor | 0 (0.0) |
| Clinical Officer | 18 (31.6) |
| Head Matron | 12 (21.1) |
| Bedside Nurse | 19 (33.3) |
| Other* | 8 (14.0) |
| **Prior Specialty Training in Pediatrics** | 13 (22.8) |
| **Number of Consultations with Children per day, median (IQR)** | 12 (6, 20) |
| **Years of Clinical Practice, median (IQR)** | 11 (6, 15) |
| **Time Practicing at Healthcare Facility** |  |
| 0-6 months | 13 (22.8) |
| 7-11 months | 11 (19.3) |
| 1-3 years | 16 (28.1) |
| 3-5 years | 8 (14.0) |
| >5 years | 9 (15.8) |
| **Clinic Location** |  |
| Peri-Urban | 30 (52.6) |
| Rural | 27 (47.4) |
| **Proportion of the respondent's clinical consultations for children under five years** |  |
| 1-5% | 9 (15.8) |
| 6-10% | 6 (10.5) |
| 11-20% | 6 (10.5) |
| 21-40% | 10 (17.5) |
| 41-60% | 16 (28.1) |
| 61-80% | 7 (12.3) |
| 81-100% | 3 (5.3) |
| **Proportion of respondent's patients (all) with Chronic Medical Problems** |  |
| 1-5% | 39 (68.4) |
| 6-10% | 10 (17.5) |
| 11-20% | 4 (7.0) |
| 21-40% | 2 (3.5) |
| 41-60% | 0 (0.0) |
| 61-80% | 1 (1.8) |
| 81-100% | 1 (1.8) |

\*Other included laboratory tech (n=1), HRIO (n=1), counselor (n=2), community health promoter (n=2) and nutritionist (n=1).

**Table 2.**
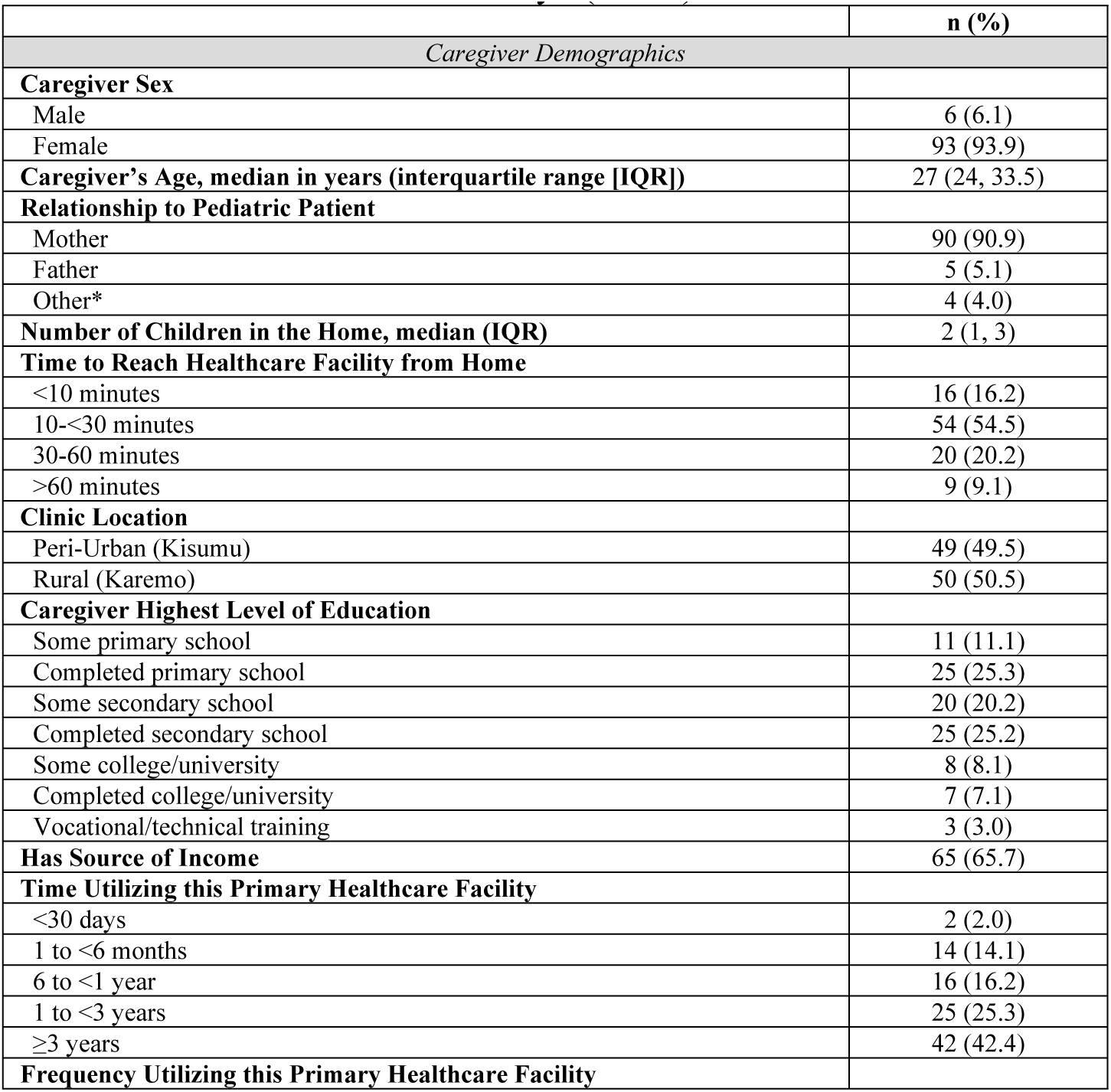
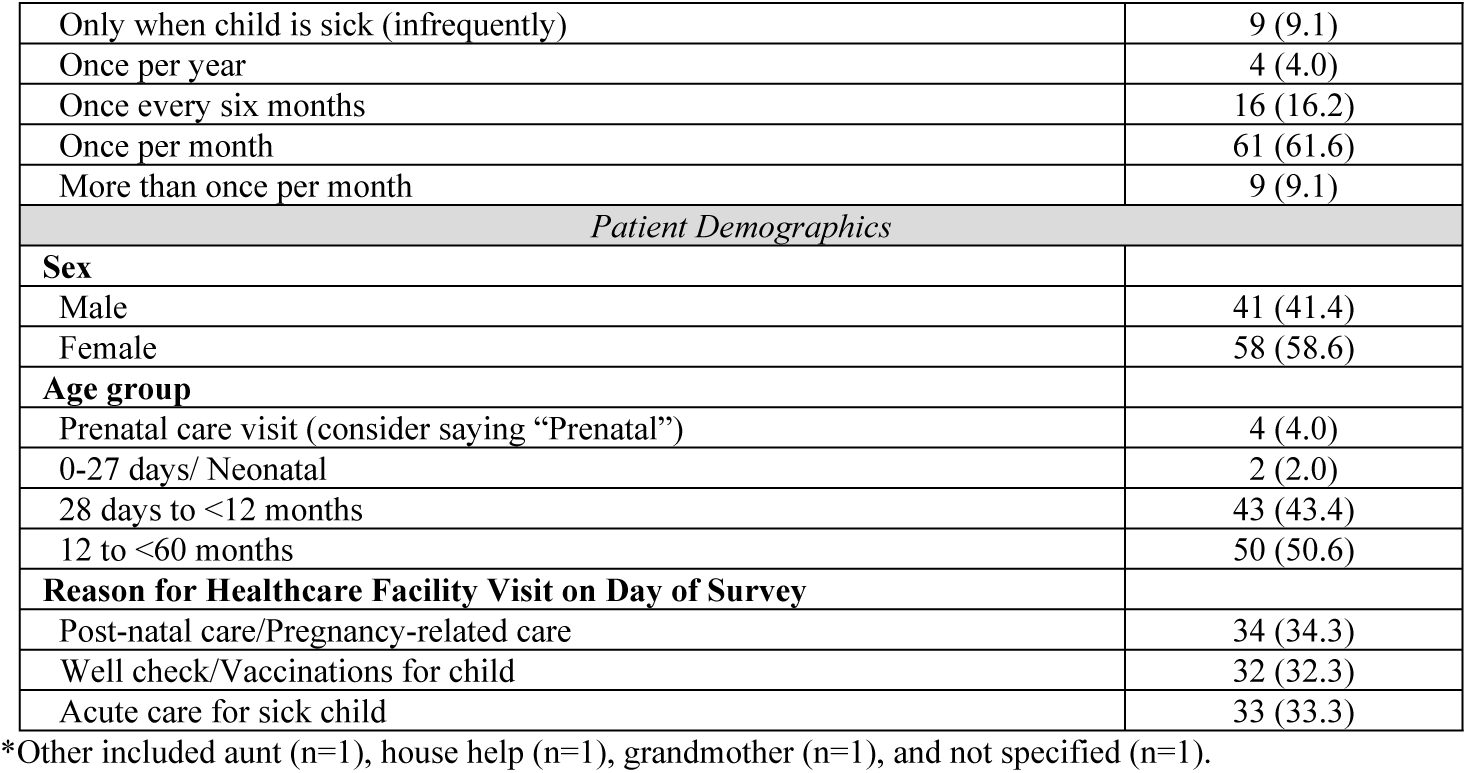
Demographics of caregivers and pediatric patients surveyed at the included primary healthcare facilities in Western Kenya (N=99)

Among the 99 surveyed caregivers, the majority were female (93.9%) and most were mothers (90.9%) with a median age of 27 years (IQR 24, 33.5 years; Table 2). Most (61.6%) caregivers reported visiting the primary healthcare facility once per month. About half of the pediatric patients (50.6%) accompanying responding caregivers were aged 12-59 months and half were younger.

### Availability of primary healthcare services

Although nearly half (47.4%) of surveyed caregivers reported that they “definitely did not wait more than 30 minutes” to be seen when in the primary healthcare facility, a notable proportion (19.3%) reported that they experienced “long wait times” (Figure 1). A majority of caregivers (98.2%) reported that they were “definitely able to see a healthcare provider on the same day for an illness when the facility was open”. Similarly, 93.0% of caregivers reportedly found it “easy to make appointments” with the primary healthcare facility. In response to a question about the “ease of receiving consultations for clinical concerns after hours and at night”, caregivers’ responses were divided (42.1% “definitely found it easy” and 36.8% “definitely did not”). In addition, 80.7% reported “definitely being able to receive advice from doctors on phone when the health facility is open” and 70.2% reported that they were able to reach a doctor on phone when the facility is closed.

**Figure 1.**
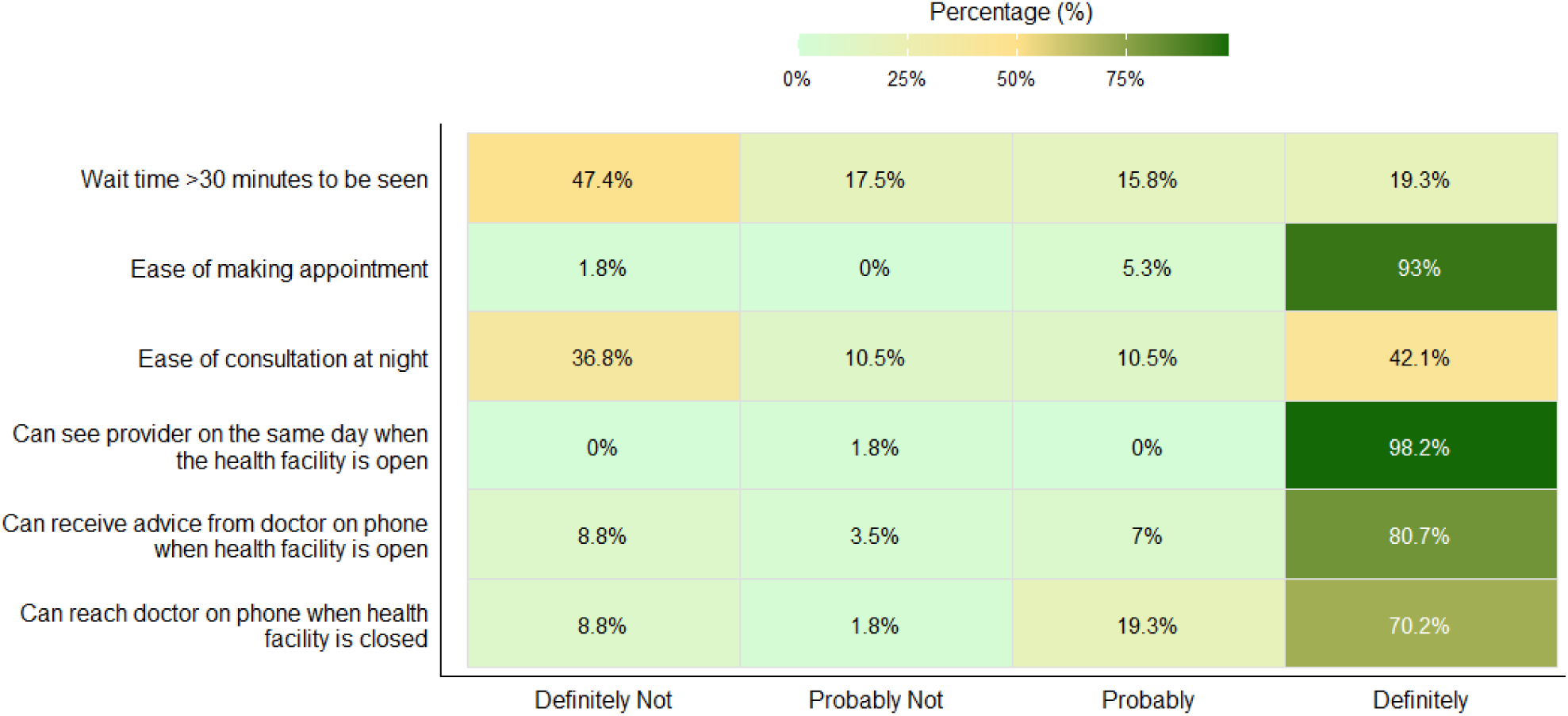
Availability of primary healthcare services according to pregnant mothers and caregivers of young children (n=99)

Many caregivers perceived a lack of after-hours primary healthcare services, particularly for same-day weekend and overnight services (Supplemental Figure 1). A smaller proportion of caregivers, 6.1% and 24.2%, believed these services were “probably” or “definitely” accessible, respectively.

In qualitative responses, caregivers reported that “none of the public primary healthcare facilities offered services over the weekend”. In contrast, they reported that private primary healthcare facilities provided 24-hour services and had more services such as specialist care and a functioning operating theater. Healthcare providers from private health facilities noted challenges with receiving payment for services following recent legislative updates that changed healthcare payment processes in Kenya. Healthcare providers also reported that healthcare services are often negatively affected by inadequate infrastructure, inadequate staffing, and lack of drugs and laboratory tests. Unlike Siaya public health facilities, respondents at public primary healthcare facilities in Kisumu reported that the maternity unit functions continuously, including at night.

> *No maternity wing but a small room for delivery where more than one woman cannot be handled. This being a small facility that should operate up to 5pm, the clinician is at times forced to work at night when there are emergencies. -Healthcare provider, Siaya*

### Comprehensiveness of Services Available

Pregnancy care (97.0%) and health talks during pregnancy (79.8%) were perceived by caregivers to be widely available (Supplemental Table 1). Delivery services (64.6%) and postpartum depression support (61.6%) were perceived to be less consistently available. Well-baby visits (97.0%) were perceived to be available by nearly all respondents, but breastfeeding support (63.6%) was perceived to be offered by fewer respondents. Routine vaccinations (94.9%), child check-ups (93.9%), treatment for diarrheal illnesses (93.9%), and malaria testing (94.9%) were perceived to be available by nearly all respondents. Screening for malnutrition was perceived to be available by 73.7% caregivers. However, fewer caregivers reported that other services such as counseling for mental health plastering for fractures were available in primary healthcare facilities. Caregivers perceived that laboratory (86.9%), pharmacy services (83.8%), and referrals (79.8%) were accessible but that transport to other healthcare facilities was usually not available (23.2%).

### Perspectives on continuity of clinical care among healthcare providers

Only 31.6% of healthcare providers reported they “definitely” knew their patients’ lives and 43.9% indicated they “probably” had this understanding (Figure 2). Nearly eight out of every ten surveyed healthcare providers (79%) reported knowing their patients’ medical history, with one-third (33.3%) expressing “definite certainty” and nearly half (45.6%) indicating “probable knowledge”. Healthcare providers’ perception of their overall knowledge of patients’ household members was more heterogeneous (17.5% reported “definitely not”, 22.8% “probably not”, 24.6%

**Figure 2.**
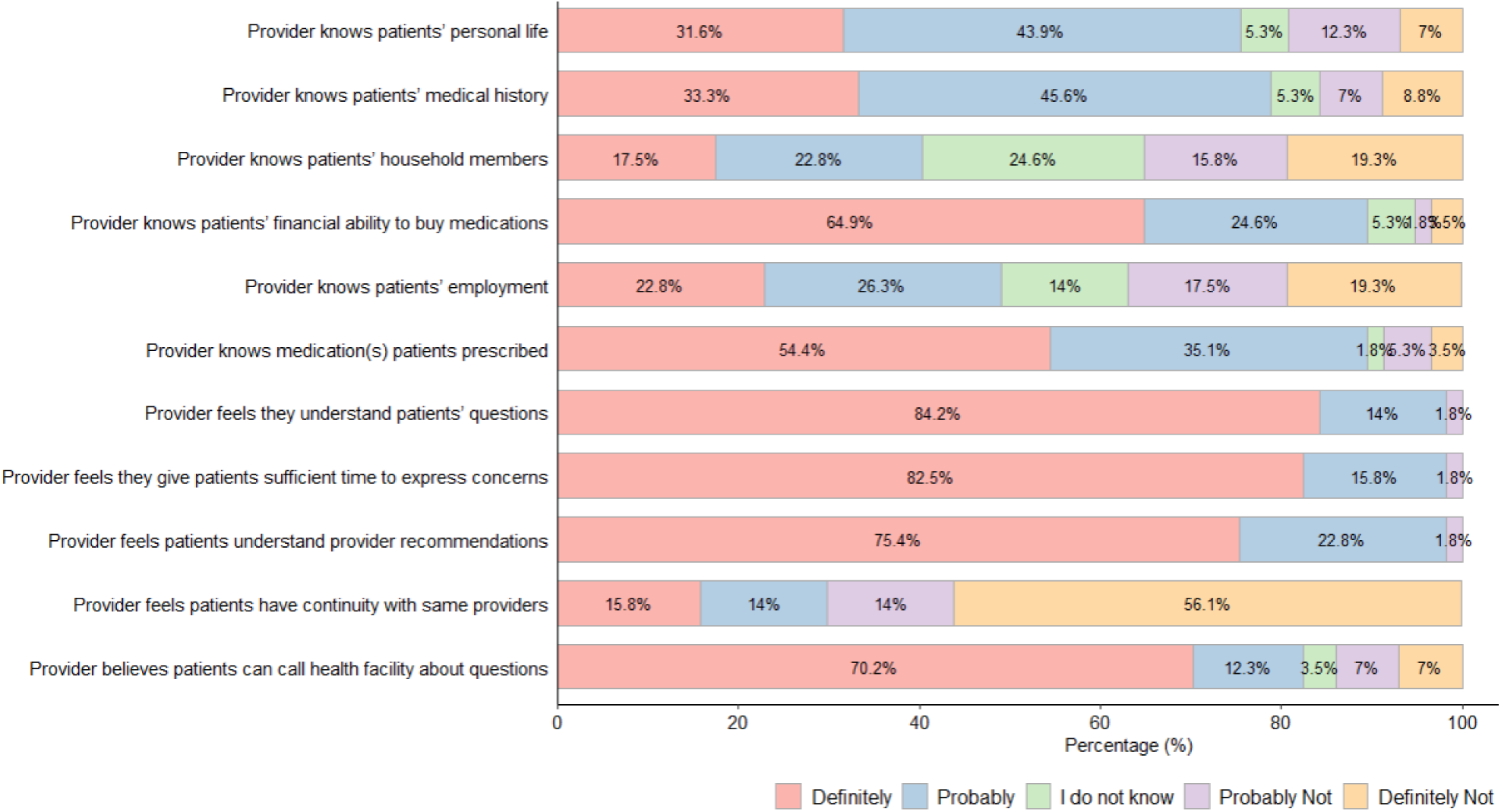
Perspectives on continuity of clinical care among healthcare providers (n=57)

“I do not know”, 15.6% “probably”, and 19.3% “definitely”). Nearly two-thirds of providers (64.9%) were “definitively confident in their knowledge of patients’ financial ability to buy medications” and 24.6% expressed “probable awareness” of the same. Providers also expressed a limited awareness of patients’ employment status with 22.8% indicating “definitive knowledge”. Slightly more than half (54.4%) expressed familiarity with medications that patients are taking.

In qualitative responses, healthcare providers indicated that primary healthcare facilities have made additional efforts to lessen the financial burden that is associated with healthcare for caregivers. Some healthcare providers reported that such efforts included designing payment plans for their patients. However, despite these efforts, healthcare providers reported that caregivers still delay seeking timely care for their children:

> *We have a designed payment plan for our patients… Some mothers bring the children to the hospital when the situation is worse…they don’t seek health care in good time. - Healthcare provider, Kisumu.*

Regarding continuity and access to health services at primary healthcare facilities, more than half of healthcare providers (56.1%) believed that patients do not have continuity with the same provider for each visit (Figure 2). Many (70.2%) expressed definite belief in patients’ ability to call the health facility with questions.

### Caregivers’ perspectives of primary healthcare facility knowledge of caregivers and child

Over half (62.6%) of caregivers reported that health facility providers “care for all the caregivers’ children” (Figure 3). There was less uniformity regarding caregivers’ confidence that the primary healthcare facility had knowledge of who lived with the child. Only 45.5% of caregivers reported that the primary healthcare facility “knows the child at a personal level”. Half of the caregivers believed that the primary healthcare facility knows the caregiver’s employment.

**Figure 3.**
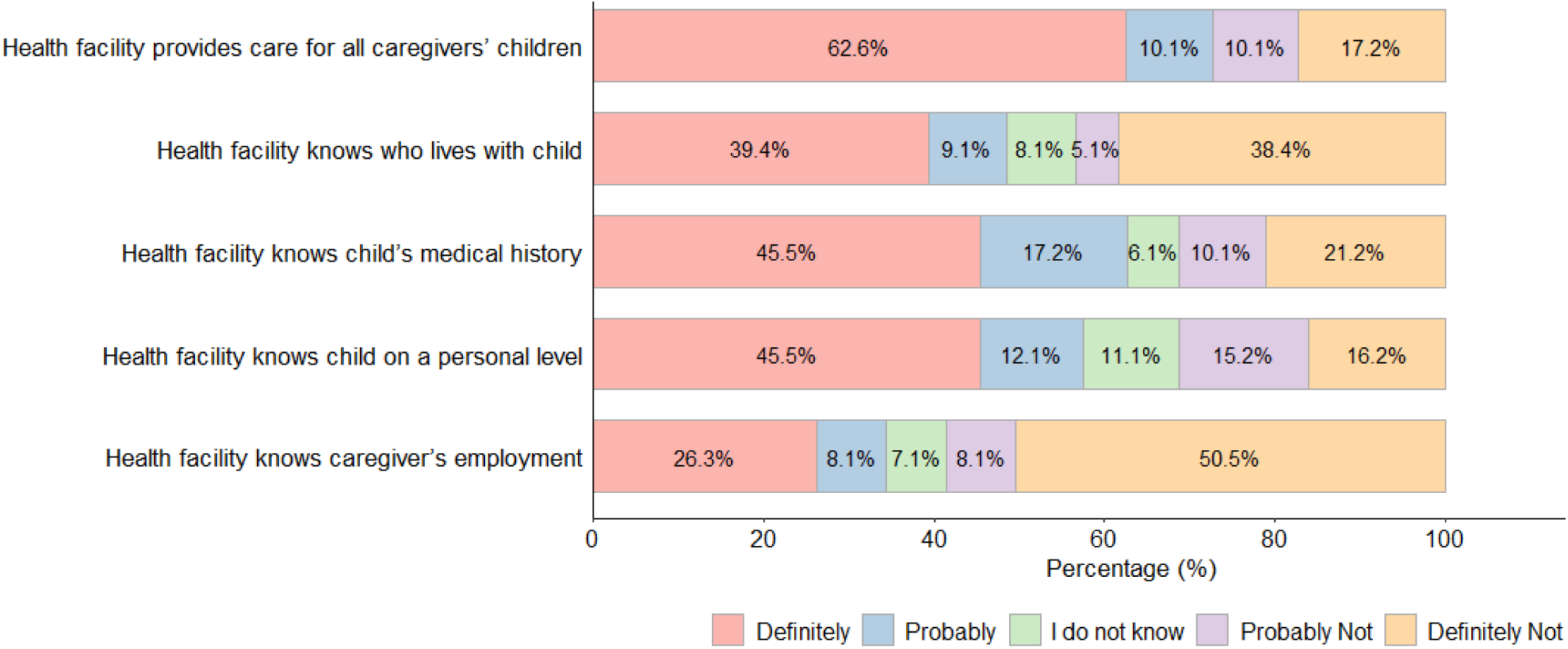
Caregivers’ perspectives on health facility knowledge of caregiver and child (n=99)

### Healthcare providers’ perception of referral systems

Most healthcare providers reported that referral letters to specialists, when indicated, were commonly provided (91.2%), yet only 45.6% reported that transportation was provided by the healthcare facility for patients to get to those specialty services.

### Caregivers’ perception of referral systems

Only 22 (22.2%) caregivers reported having been referred to another healthcare facility in the past, out of whom 10 (45.5%) reported that the primary healthcare facility followed up to discuss the referral visit and 12 (54.5%) reported that the primary healthcare facility seemed interested in the quality of care received from that specialist or hospital.

### Perspectives on community orientation of healthcare services among healthcare providers

Most healthcare providers (77.2%) reported that they consider patients’ opinions and ideas to inform healthcare provision (Figure 4). Additionally, 66.7% of healthcare providers reported considering patients’ language when informing the approach to healthcare provision. In contrast, only 43.9% of healthcare providers took “special beliefs about healthcare into account”.

**Figure 4.**
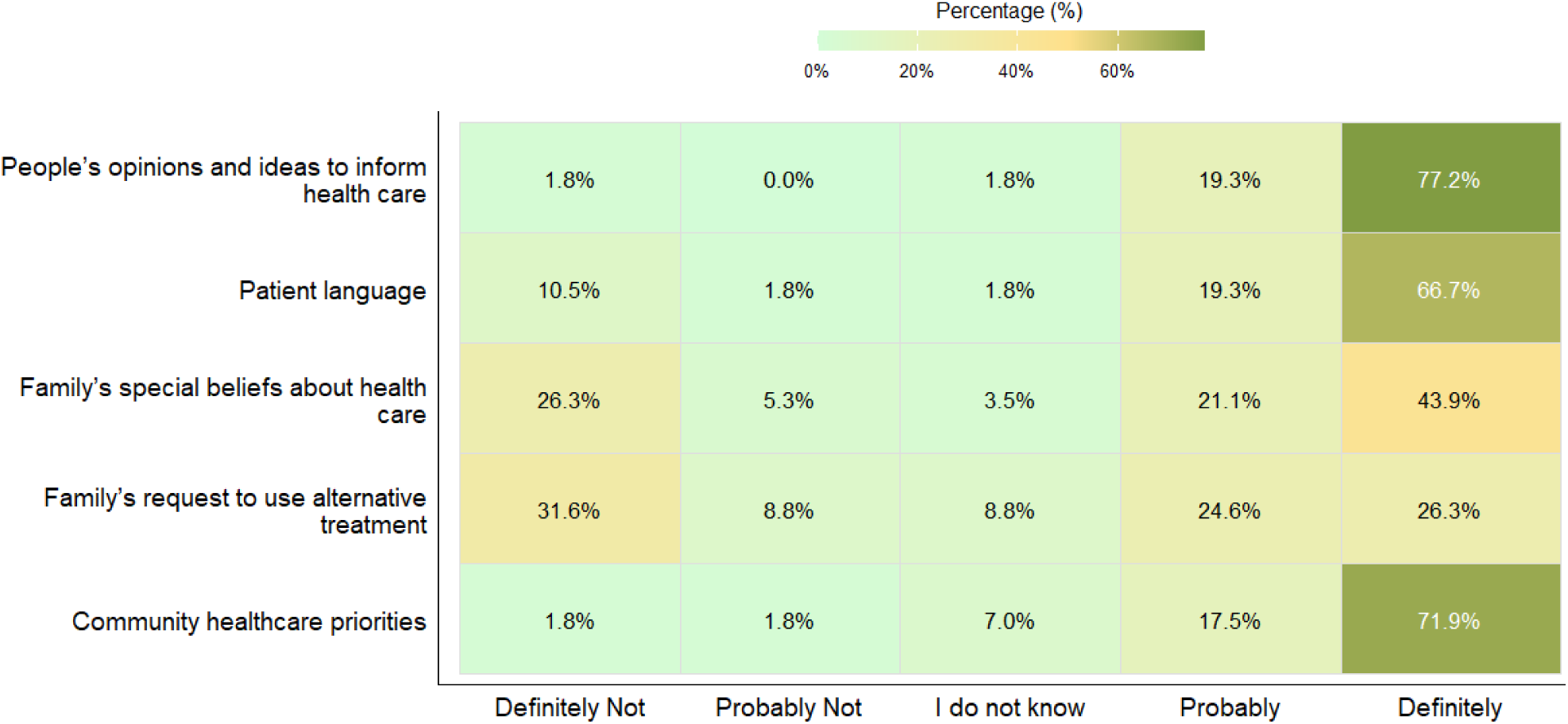
Perspectives on community orientation of primary healthcare services among healthcare providers (n=57)

**Figure 5.**
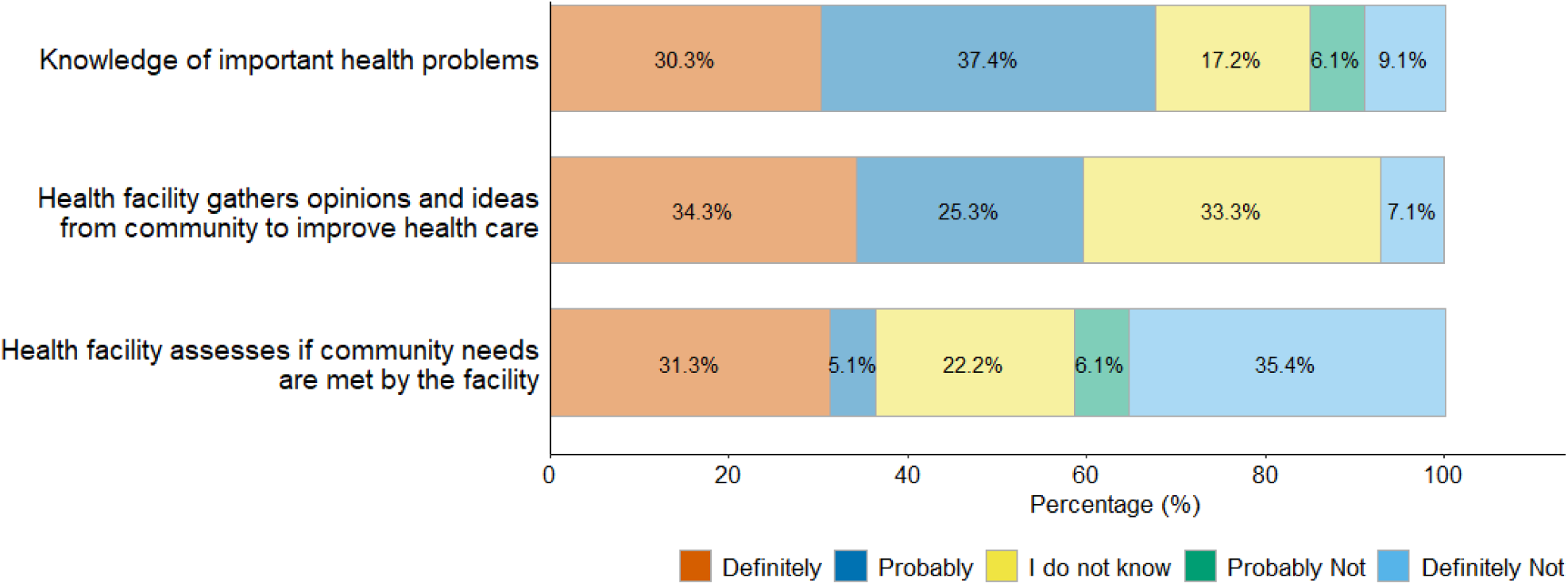
Caregivers’ perspectives on community orientation of primary healthcare providers (n=99)

Healthcare providers had varied perceptions about caregivers’ request to use alternative treatment: 23.6% said they “definitely take it into account”, 31.6% reported “definitely not”, and 24.6% reported “probably would take it into account”. A sizable proportion (71.9%) said that they would consider community health priorities.

### Caregivers’ perspectives on community orientation of primary healthcare providers

Most caregivers (67.7%) reported that the health facility is “aware of their important health problems” (Figure 6). However, when asked whether the facility assesses whether community needs are being met, responses were mixed: 31.3% said “definitely,” 35.4% said “definitely not,” and 22.2% indicated they “did not know”. Similarly, caregivers expressed varied views on whether the facility actively gathers opinions and ideas from the community to improve healthcare services. While 34.3% believed this “definitely happens”, 25.3% disagreed, and a significant proportion (33.3%) “did not know”.

## Discussion

Our study of healthcare providers and caregivers of pediatric patients and pregnant women in 20 primary healthcare facilities in Western Kenya revealed varied level of availability and quality of primary healthcare services. Specifically, there was limited access to primary healthcare services outside the standard weekday hours, which aligns with findings from previous study in urban regions in Kenya.(24) However, private healthcare facilities were perceived to be more accessible. Yet, caregivers reported that these services may not be widely available due to associated costs in private healthcare facilities.(25) All primary healthcare facilities included in this study reported providing antenatal and malnutrition screening for children; however, fewer provided newborn care services and postpartum clinical care. As maternal and newborn deaths are a particularly sensitive indicator of health system quality, such services may be targeted for future efforts to strengthen health systems in Western Kenya.(26,27)

In our study, healthcare providers perceived wide availability of health services during official working hours. However, our findings suggest that challenges remain in areas related to timeliness and after-hours care in primary healthcare facilities in Western Kenya. For instance, a significant number of providers reported that making appointments is easy and nearly all affirmed same-day consultations when the facility is open; however, long wait times were reportedly common. Primary healthcare facility responsiveness to patient needs is an important aspect of healthcare,(28,29) yet reports indicate it remains low in nearby countries in East Africa.(30)

Most healthcare providers reported that referral letters to specialists were commonly provided. However, in very few instances transport was available from the primary healthcare facility to enable these referrals. These findings are consistent with those in a previous study that reported that one-third of pediatric patients who died were seen in primary healthcare facilities yet were not referred, often due to challenges in transportation.(10) Evidence from a scoping review suggests a strong relationship between primary healthcare facilities and hospitals, effective gatekeeping management of referrals, and patient-centered communication may enhance the efficiency of the referral system.(31) However, some healthcare facilities lacked communication mechanisms to ensure coordination between primary health facilities and referral hospitals.

Overall, these findings highlight how deficiencies in both structural support such as transport and relational processes such as communication and coordination hinder the effectiveness of referral systems, despite providers’ willingness to refer patients for higher-level care.

Our findings suggest that although healthcare providers had a strong understanding of patients’ medical histories, their understanding of patients’ broader social circumstances was uneven. Such understanding is crucial as social factors such as financial capacity to purchase medications may influence treatment adherence and overall health outcomes.(32) A previous systematic review highlighted potential benefits to healthcare providers’ knowledge of patients’ social circumstances, which includes improved healthcare equity and higher quality healthcare service provision.(33) Despite documented benefits, there remains limited empirical research exploring the extent to which providers integrate social context into patient care, particularly in low- and middle-income country settings. In addition, our results demonstrate healthcare providers in Western Kenya were relatively unfamiliar with the medications taken by their patients, pointing to possible communication gaps or fragmented record-keeping that could negatively affect continuity of care.(32)

Most caregivers reported that healthcare facilities were aware of their important health problems. However, they were unsure of whether healthcare facilities met community needs and whether opinions about healthcare needs were met. In contrast, many healthcare providers reported considering patients’ needs and requests when providing clinical care. The findings indicate a possible divergence between healthcare provider self-assessment and caregiver perceptions. Past studies have reported inadequate accommodation of patients’ needs, with many health systems continuing to prioritize acute illness management while neglecting the ongoing requirements of chronic disease care.(32)

### Limitations

The findings from this facility-based PCAT study may not be fully generalizable to other healthcare facilities throughout Kenya. The specific challenges in the provision of primary healthcare facility-based clinical care for pregnant women and children aged <5 years may not be representative of challenges faced by other facilities in the catchment area.

Although only 56% of caregivers approached participated in the study, the sample included diverse participants across key demographic characteristics. Therefore, while some degree of selection bias cannot be ruled out, the findings are likely to be broadly representative of the study population. In addition, caregivers of children may have additional insights regarding reasons for gaps in clinical care that will not be captured with the current approach, which may warrant future investigation. The PCAT measures perceptions of healthcare and not delivered healthcare. Lastly, although our team has fluency in English, Dholuo and Kiswahili and translated the questionnaires, the PCAT was translated from English to Kiswahili or Dholuo, which may have resulted in some elements not understood as originally intended.

## Conclusions

Although services were generally accessible for pediatric and antenatal care, postnatal care was not widely available in primary healthcare facilities in Western Kenya. Healthcare providers’ knowledge about patient’s beliefs was low, and the referral system was undermined with coordination gaps and transportation challenges. The quality and availability of clinical care at primary healthcare facilities may be improved by improving continuity of care, coordination of care and comprehensiveness of services (both available and provided). Urgent attention needs to be given to strengthening postnatal service delivery and improving referral system coordination.

## Data Availability

All data will be made available.

## Acknowledgements

The authors are immensely grateful to all the participants who took part in this study. We thank the CHAMPS program Kenya for supporting this study. We would like to acknowledge the support of CHAMPS program staff in collecting this data and Emory/CHAMPS program office for technical support.

## Abbreviations

PCAT: Primary Care Assessment Tool
CHAMPS: Child Health and Mortality Prevention Surveillance

---

**Supplemental Figure 1.**
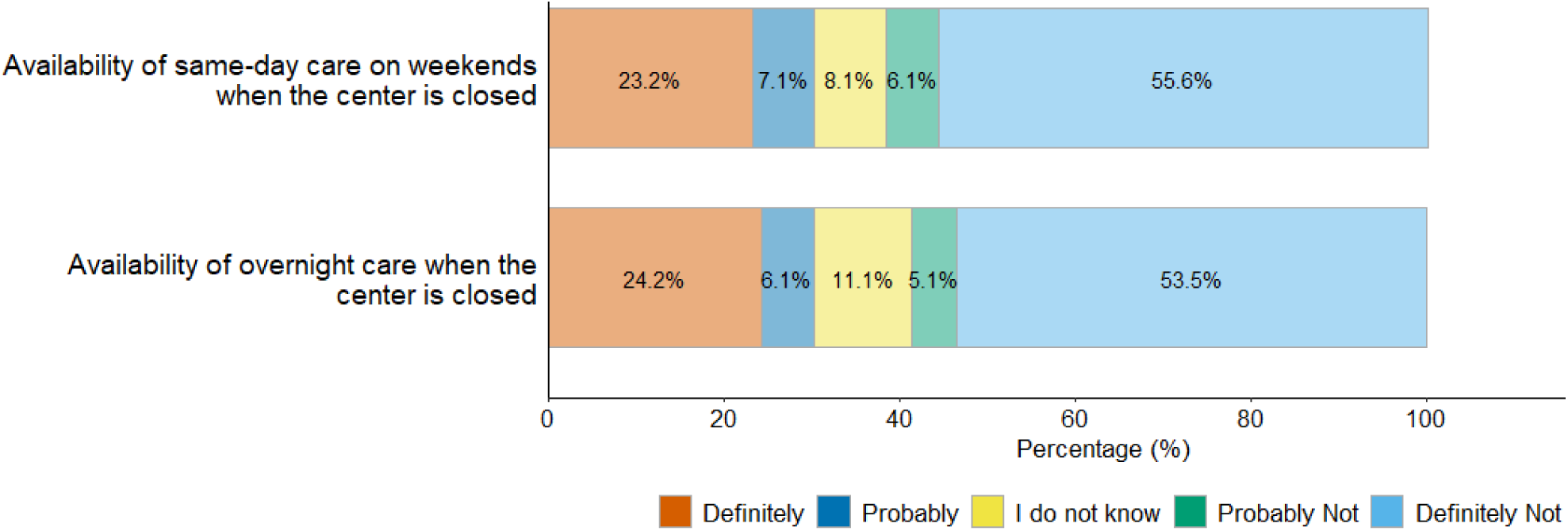
Caregiver perceptions of availability of primary healthcare services availability after hours.

**Supplemental Table 1.** Caregiver perspective on comprehensiveness of services available (n=99)

| Available at Healthcare Facility | n (%) |
| --- | --- |
| *Pregnancy Care* | 96 (97.0) |
| Health Talks During Pregnancy | 79 (79.8) |
| Delivery | 64 (64.6) |
| Postpartum Counselling | 61 (61.6) |
| *Newborn Care* |  |
| Well Baby Visits | 96 (97.0) |
| Breastfeeding Support | 63 (63.6) |
| *Child Care* |  |
| Routine Vaccinations | 94 (94.9) |
| Routine Child Check-Up Visits | 93 (93.9) |
| Screening for Malnutrition | 73 (73.7) |
| Counseling for Mental Health Challenges | 39 (39.4) |
| Treatment for Diarrheal Illnesses | 93 (93.9) |
| Malaria Testing | 94 (94.9) |
| Plastering for Fractures | 28 (28.3) |
| Treatment for Tuberculosis | 55 (55.6) |
| Testing and Treatment for HIV | 90 (90.9) |
| *Laboratory Services* | 86 (86.9) |
| *Pharmacy Services* | 83 (83.8) |
| *Referrals to Other Healthcare Services* | 79 (79.8) |
| *Transport to Other Healthcare Services* | 23 (23.2) |

